# Causal factors affecting gross motor function in children diagnosed with cerebral palsy

**DOI:** 10.1101/2020.10.26.20217232

**Authors:** Bruce A. MacWilliams, Sarada Prasad, Amy L. Shuckra, Michael H. Schwartz

## Abstract

Cerebral palsy (CP) is a complex neuromuscular condition which may negatively impact gross motor function. Children diagnosed with CP often exhibit spasticity, weakness, reduced motor control, contracture, and bony malalignment. Despite many previous association studies, the causal impact of these impairments on motor function is unknown. In this study, we propose a causal model for motor function as measured by the 66-item Gross Motor Function Measure (GMFM-66), and estimate the direct and total effect sizes of these common impairments using linear regression based on covariate adjustment sets implied by the causal model. We evaluated 300/314 consecutive subjects with cerebral palsy who underwent routine clinical gait analysis. The largest effect sizes, as measured by standardized regression coefficients (standard error), were for static motor control (direct = 0.35 (0.04), total = = 0.40 (0.04)) and dynamic motor control (direct = 0.26 (0.04), total = 0.31 (0.04)), followed by strength (direct = 0.23 (0.04), total = 0.26 (0.04)). The next largest effect was found for gait deviations (direct = total = 0.15 (0.04)). In contrast, common treatment targets, such as spasticity (direct = 0.05 (0.03), total = 0.08 (0.03)) and orthopedic deformity (direct = 0.00 (0.03) to 0.08 (0.03), total = −0.01 (0.03) to 0.11 (0.03)), had relatively small effects. We also show that effect sizes estimated from bivariate models, which fail to appropriately adjust for other causal factors dramatically overestimate the total effect of spasticity (510%), strength (271%), and orthopedic deformity (192% to -2017%). Understanding the relative influences of impairments on gross motor function will allow clinicians to direct treatments at those impairments with the greatest influence on gross motor function and provide realistic expectations of the anticipated functional changes.

## Introduction

### Gross Motor Function in Cerebral Palsy

Gross motor function in children directly influences quality of life indicators such as activity and participation^1–3^. Children diagnosed with cerebral palsy (CP) often have limitations in their gross motor abilities^4^. An important goal of treatment for these children is to optimize functional abilities. However, knowing what to treat is not trivial, since the link between neurological and musculoskeletal factors and gross motor skills is poorly understood. There is little evidence that common treatments for the CP child effectively improve function. Significant improvements in gross motor function have not been found with single event multilevel surgery^5–7^. Selective dorsal rhizotomy may result in small functional increases at short- and mid-term, but these are not maintained at long term follow-up^8–12^. Likewise, evidence of functional gains from physical therapy treatments are limited to short-term changes^13–15^. Knowing the relative influences of impairments on gross motor function can allow clinicians to direct treatments at those impairments with the greatest impact on this important outcome domain.

Cerebral palsy is a complex neurological condition. Damage to the developing brain impairs motor control and may induce spasticity, leading to reduced strength and abnormal musculoskeletal loading, further leading to joint contractures and bony deformities. Although the injury is non-progressive, secondary effects can continue to worsen with maturation. Function in this population is most commonly measured using the Gross Motor Function Measure (GMFM), a task-based standardized test consisting of five domains (A. Lying and Rolling, B. Sitting, C. Crawling and Kneeling, D. Standing, and E. Walking, Running and Jumping) that rate a patient’s level of function^16^. The GMFM may be reported using the full 88-item score (GMFM-88), a 66-item score (GMFM-66)^17^, or individual domain scores. The GMFM has been shown to be related to measures of mobility, self-care, and social function^18,19^. For ambulatory children, the standing (GMFM-D) and walking, running, jumping (GMFM-E) domain scores are highly relevant to mobility.

### Association Studies of Function

Much of the literature investigating impairments and function consists of reports examining the effects of a few chosen variables (*e*.*g*. strength of a single joint). Several such studies have investigated relationships between impairments and gross motor function using bivariate correlation or regression: analysis of one independent variable of cause (*e*.*g*. spasticity, strength) to one dependent variable of effect (*e*.*g*. gait, GMFM)^20–23^. This approach is insufficient since it ignores the dependent effects of other influential variables. These studies focus on associations, with little or no formal discussion of causal mechanisms. For example, Shin *et al*. evaluated bivariate correlations between isometric hip and knee strength in 24 pediatric subjects with CP and an extensive list of 30 kinematic and kinetic parameters, GMFM-D, and GMFM-E. Correlations with GMFM domain scores ranged from *r*=0.06-0.30, and none were significant. Other primary (*e*.*g*. spasticity, motor control) and secondary (*e*.*g*. contracture, bony torsion) were not tested, and each strength measure was considered in isolation^22^.

When multivariate relationships have been explored, it has been done with limited variable sets, small numbers of participants, and without explicitly proposing and testing causal relationships. Eek and Beckung examined the relationships between hip, knee and ankle strength and function in 55 children with CP. No other variables were considered. In contrast to Shin, significant bivariate correlations were found ranging from *r*=0.59-0.80 between strength and GMFM-66. The multivariate combination most strongly correlated to GMFM-66 included ankle plantarflexors and hip flexors, yielding an *r*^*2*^=0.73^24^. Curtis *et al*. used a hypothesized multivariate model to investigate the influence of trunk control on GMFM-88 with age and neuromotor impairment type as covariates in 92 subjects^25^. Multivariate correlations ranged from *r*^*2*^=0.49-0.51 with trunk control accounting for 38-40% of the variance.

While previous studies have identified many factors *associated* with gross motor function, the utility of these models for guiding intervention is limited since pernicious effects like confounding, mediation, and opening of non-causal pathways is possible^26^. Remarkably, Eek and Beckung reported that just two muscle groups account for 73% of the variance in GMFM-66^24^. In addition to straining credulity, a problem with this finding is that strength is analyzed in isolation, without consideration for the causal influences of other variables. On the surface, Eek and Beckung’s results would suggest strengthening as a highly effective therapy. However, neurological impairments do not occur in isolation. If weakness is influenced by poor motor control, age, or overall severity, the effects of strengthening might be smaller than estimated. In what follows we will show that proper accounting for covariates reduces the apparent importance implied by non-causal analysis of many individual clinical factors.

### Explicit Causal Approach

Behind most, if not all, association studies lurks an implicit causal hypothesis. It is unlikely that Eek and Beckung were simply trying to demonstrate an association between strength and GMFM-66. Rather, their study almost certainly reflects an assumption that strength is an important *cause* of GMFM-66. This assumption is entirely plausible. In fact, we will demonstrate below that strength *is* an important causal factor, though substantially smaller than bivariate estimates would suggest. However, if causal claims are going to be made they should be explicit and they should be tested.

An explicit causal model provides a set of conditions that assess the plausibility of the model. Once plausibility is established, a casual model produces covariate adjustments necessary for statistical modeling. There is limited use of causal modeling in the study of CP. Kim and Park proposed a simple causal model to examine the effects of spasticity and strength on GMFM-88 and functional outcomes as measured by the Pediatric Evaluation of Disability Inventory (PEDI)^27^. Only GMFM-88 had significant direct effects on PEDI, whereas spasticity and strength had indirect effects mediated by GMFM-88. The authors noted that the model lacked other potentially important variables such as orthopedic deformities.

Instrumented three-dimensional gait analysis (gait analysis) is a common diagnostic and treatment-planning tool used in the assessment of children with CP. Standard gait analysis protocols include direct and indirect measures of neurological and orthopedic impairments that may influence function. In this study, we will propose and test a causal model for GMFM-66 that examines the influence of key neurological and orthopedic impairments commonly measured during gait analysis. Compared to Kim and Park’s model, the proposed model will add measures of orthopedic deformity, gait quality, and motor control, including a recently developed measure of dynamic motor control^28^. Finally, in addition to reporting results, we will test the plausibility of the model. Understanding the causal factors influencing function can better educate patients, families, and providers about realistic goals for treatments.

## Results

Participant demographics reflect the population of CP patients referred for clinical gait analysis (Table 1). We screened 314 individuals to reach the final cohort of 300 participants. We excluded 14 subjects (4%) who had missing data (6 missing selective control assessment of the lower extremity – SCALE^29^ scores, 13 missing strength scores). Three of the excluded subjects had missing data due to visit time constraints; the other 11 were unable to complete testing due to an inability to follow directions. Of these 11, 5 were age 6 years old or younger.

**Table 1.** Participant Characteristics.

|  | GMFCS I | GMFCS II | GMFCS III |
| --- | --- | --- | --- |
| N | 115 | 132 | 53 |
| CP Sub-type (N) |  |  |  |
| diparesis, paraparesis | 30 | 95 | 47 |
| hemiparesis | 85 | 32 | 0 |
| quadraparesis | 0 | 5 | 6 |
| Gender = Male/Female (N) | 52/63 | 75/57 | 34/19 |
| Age (years) | 10.5 (3.52) | 11.6 (3.46) | 10.9 (3.25) |
| Walk-DMC [100 (10)] | 91.0 (9.99) | 79.7 (12.16) | 66.5 (9.83) |
| GDI [100 (10)] | 82.1 (10.69) | 70.9 (9.44) | 62.6 (8.47) |
| SCALE [10] | 6.8 (1.35) | 5.2 (1.43) | 3.0 (1.42) |
| Spasticity [0] | 1.6 (0.91) | 2.1 (1.12) | 2.9 (1.10) |
| Strength [5] | 4.6 (0.30) | 4.1 (0.48) | 3.5 (0.61) |
| Speed (m/s) [1.2 (0.12)] | 1.1 (0.15) | 0.9 (0.19) | 0.6 (0.21) |
| GMFM-66 [100] | 86.8 (6.23) | 73.1 (7.01) | 57.0 (4.64) |
| Ankle Dors. Contracture (degrees) [0] | 15.5 (6.06) | 18.2 (8.14) | 18.6 (8.17) |
| Knee Ext. Contracture (degrees) [0] | 4.8 (4.87) | 2.3 (7.21) | -3.4 (8.75) |
| Hamstrings Contracture (degrees) [0] | 8.9 (17.37) | 19.6 (18.70) | 26.1 (18.34) |
| Hip Ext. Contracture (degrees) [0] | 0.9 (2.65) | 3.8 (5.10) | 7.6 (7.57) |
| Femoral Anteversion (degrees) [0] | 40.3 (19.81) | 43.7 (20.24) | 50.2 (21.60) |
| Tibial Torsion (degrees) [0] | 5.0 (3.60) | 5.5 (5.06) | 6.8 (5.06) |

*Note: All values given as mean (standard deviation). These are raw, untransformed values. For modeling purposes, variables were transformed so that larger values indicate less impairment, and smaller values indicate more impairment. Typically developing values are given in [mean (sd)]*

The implied independencies were tested, and all partial correlations were found to be less than 0.11 and statistically insignificant. This demonstrated that the structure of the proposed causal model was consistent with the observed data. This does not prove the proposed model is correct, but it does prove the proposed model is plausible. Standardized coefficients were computed to assess effect sizes for the various neurological and orthopedic factors (Figure 1).

**Figure 1.**
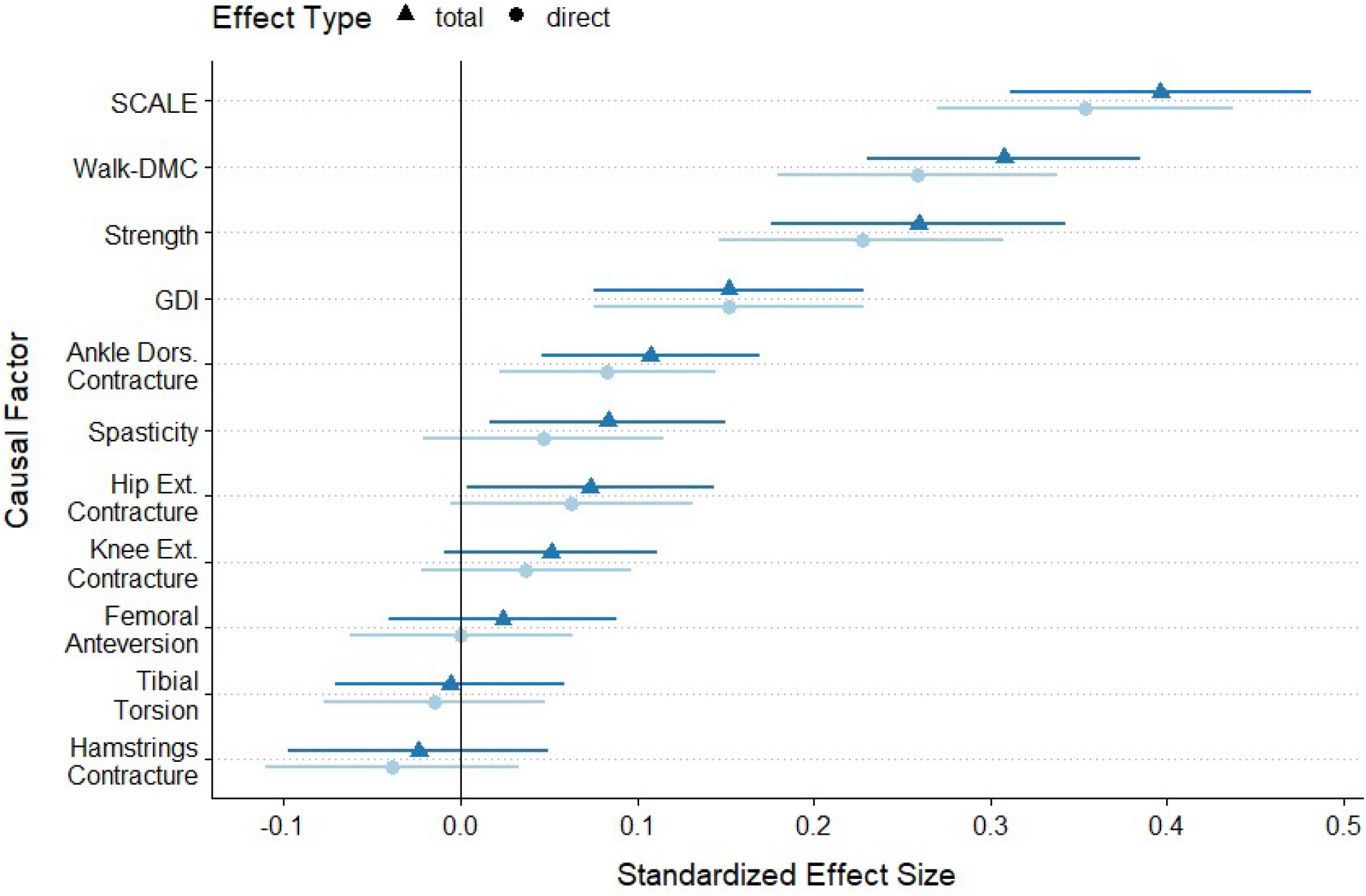
Direct (light blue) and total (dark blue) effect sizes of causal factors are expressed as the standardized regression coefficients (95% CI shown). Static and dynamic motor control (SCALE and Walk-DMC) have the two largest effects on GMFM-66, followed closely by Strength. Spasticity had a modest total effect, and a large decrease from total to direct effect size, suggesting its action is significantly mediated by other factors. From an orthopedic perspective, Ankle Dorsiflexion and Hip Extension had effect sizes similar to Spasticity, Knee Extension had a marginal effect, and other orthopedic impairments did not meaningfully influence GMFM-66. Note that due to the hypothesized causal model structure, the direct and total effects of GDI are the same. Also note that GDI is not directly manipulable by treatment, but rather it changes in response to modification of orthopedic or neurological factors.

Comparing effect sizes between the causal model and the bivariate regression models shows the important impact that causal modeling can have on apparent effect sizes (Figure 2).

**Figure 2.**
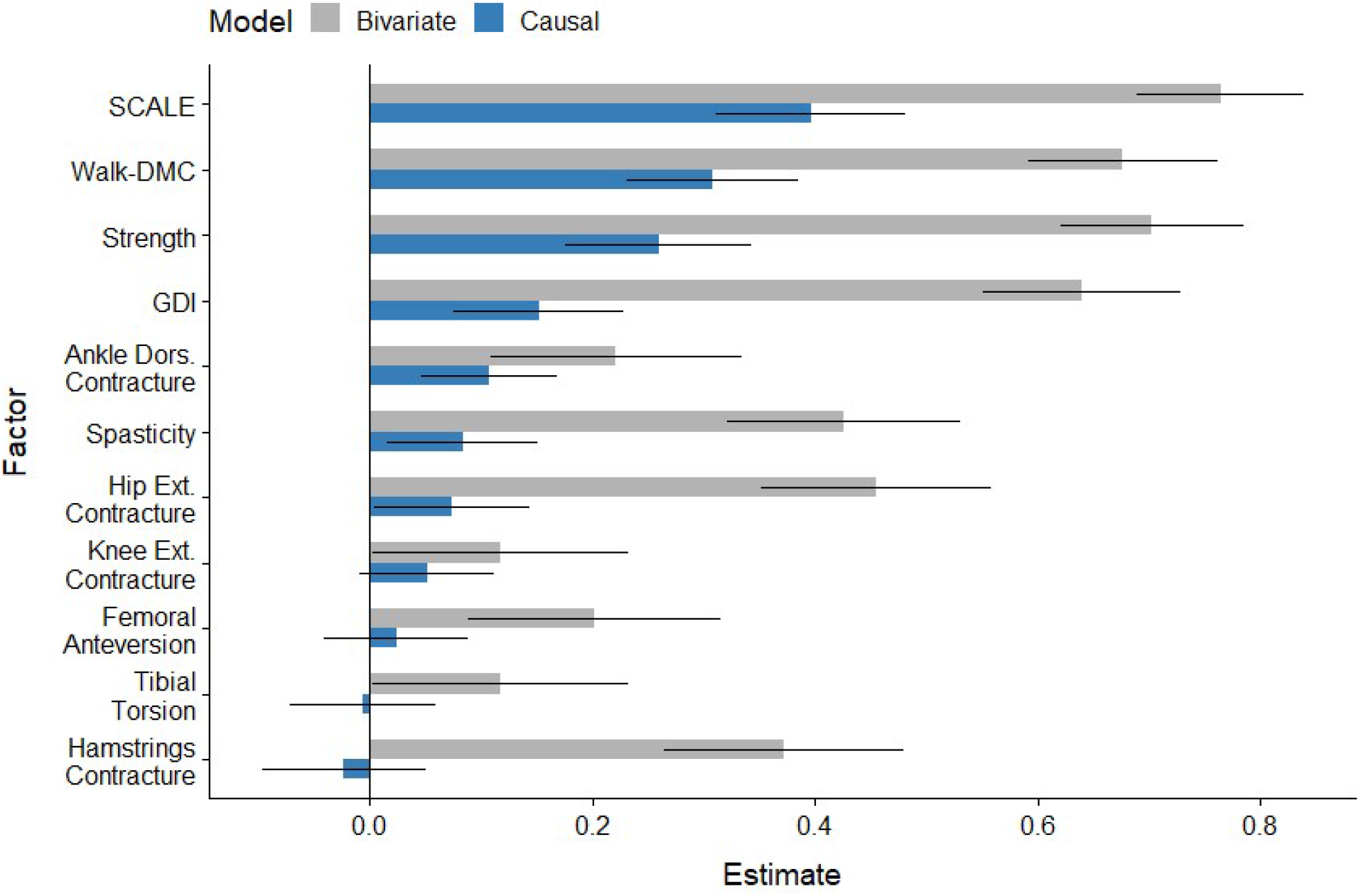
Comparison of bivariate (grey) and total causal (blue) effect sizes shows the impact that causal modeling has on the estimated importance of clinical factors. The most obvious difference is one of magnitude – where bivariate effect sizes significantly overestimate the influence of each factor. In addition, the bivariate models suggest large effects for Hip Extension, Spasticity, Hamstrings Contacture and Femoral Anteversion. After proper adjustment via causal modeling, each of these effects shrinks dramatically.

Bivariate (non-causal) effect sizes overestimated the importance of factors by 192% to -2017%, compared to their causal counterparts (Figure 2).

Coefficients for the direct linear model were estimated for all causal factors (Table 2). A 10-fold cross-validation was performed to estimate this model’s performance on out-of-sample observations. This resulted in an out-of-sample *r*^*2*^ = 0.75 and a mean absolute error of 4.9 points (Figure 3).

**Table 2.** Predictive model coefficients.

| Variable | $\beta$ | SE | | Units | Range | TD Values | Description |
| --- | --- | --- | --- | --- | --- | --- | --- |
| Intercept | 16.5 | 4.5 | <.01 | S | NA | NA | GMFM-66 linear intercept<sup>17</sup> |
| Age | 0.03 | 0.11 | .80 | Yr | NA | NA | Child's age in decimal years (e.g. 10.4 yrs) |
| SCALE | 2.2 | .27 | <.01 | S | 0-5 | 5 | SCALE total limb score bilateral average (0-10)<sup>29</sup> |
| Walk-DMC | 0.23 | 0.04 | <.01 | S | 22-100+* | 100 (10) | Dynamic motor control scaled from normal<sup>28</sup> |
| Strength | 4.8 | 0.85 | <.01 | S | 0-5 | 5 | Manual muscle grade average<sup>30</sup> |
| Spasticity | -0.52 | 0.38 | .17 | S | 0-5 | 0 | Multivariate spasticity scale<sup>34</sup> |
| GDI | 0.15 | 0.04 | <.01 | S | 0-100+* | 100 (10) | Gait deviation index<sup>31</sup> |
| Ankle Dors. Contracture | -0.14 | 0.05 | <.01 | Deg. | >=0 | 0 | Limitation in achieving 15 of dorsiflexion with knee extended<sup>32,33</sup> |
| Hip Ext. Contracture | -0.15 | 0.08 | .06 | Deg. | >=0 | 0 | Limitation in achieving neutral hip extension<sup>32,33</sup> |
| Knee Ext. Contracture | -0.09 | 0.07 | .20 | Deg. | >=0 | 0 | Limitation in achieving neutral knee extension with hip at neutral<sup>32,33</sup> |
| Hamstrings Contracture | -0.02 | 0.02 | .30 | Deg. | >=0 | 0 | Limitation in achieving neutral knee extension with hip flexed to 90° (Popliteal Angle)<sup>32,33</sup> |
| Femoral Anteversion | -0.00 | 0.02 | .98 | Deg. | >=0 | 0 | Absolute difference from normal anteversion (10°)<sup>35</sup> |
| Tibial Torsion | -0.04 | 0.08 | .65 | Deg. | >=0 | 0 | Non-weightbearing bimalleolar axis difference from normal (15°)<sup>32,33</sup> |

*S* = unitless scale variable as defined in description. *NA* = not applicable. *TD* = typically developing. *SE* = standard error. Added together, the coefficients $\beta$ multiplied by the values for a subject yield a predicted GMFM-66 score.
\*Both Walk-DMC and GDI are scaled Z-scores and can exceed the mean typical value of 100, additionally Walk-DMC has a theoretical minimum value dependent upon the scaling values.

**Figure 3.**
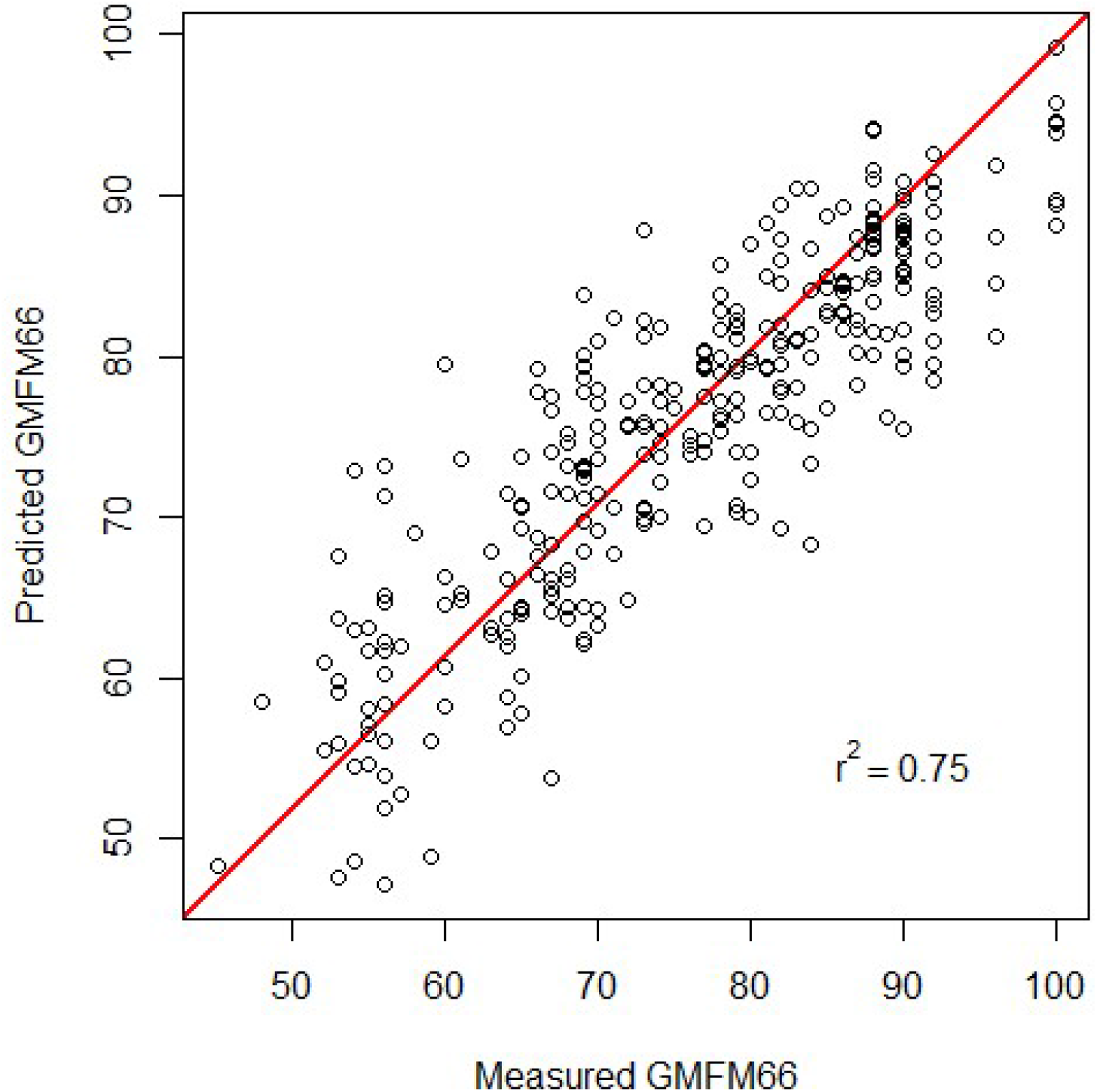
Predicted *vs*. measured GMFM-66 for the final total effects linear model. The diagonal line indicates perfect agreement. The cross-validated r^2^=0.75 and mean absolute error = 4.9 points.

## Discussion

A causal model of function in children with CP provided estimates of direct and total effect sizes for key clinical impairments, and explained 75% of the variance in the GMFM-66 scores in an out-of-sample test set (10-fold cross-validation). The model is plausible and can be evaluated using measurements collected during routine gait analysis evaluations. Static motor control, dynamic motor control, and strength have the largest total effects on GMFM-66; more than three times larger than the effect of spasticity. Gait impairments (GDI) have the next largest impact on GMFM-66. Ankle and hip contractures have small effects on GMFM-66. The causal effects of most clinical factors were found to be substantially smaller than those obtained from bivariate analysis.

Our findings have important clinical implications. First, the effects of these measures on gross motor function can guide targets for treatment and expectations regarding possible improvements. Second, the results confirm that factors explaining 75% of gross motor function can be measured during routine gait analysis. Finally, the results demonstrate the errors that can arise from naively computing bivariate relationships among clinical factors in children diagnosed with CP. The interconnectedness of neurological and musculoskeletal impairments, gait, and function in this population places a premium on employing analyses that respect the complexity of the clinical condition.

Static selective motor control, as measured by the SCALE composite limb score, had the highest effect in the total effects model. The SCALE score has previously been *associated* with GMFM- 66, and our analysis confirms that this association appears to be causal^36^. Dynamic motor control, as measured by Walk-DMC^28^, had a large effect size, even after controlling for static motor control. This suggests that the neurological pathways employed during quasi-static tasks and those used during gait are not the same. It is important to note that, in our proposed model, the impairments in static and dynamic motor control share a common cause and are hence correlated (Walk-DMC ← Injury → SCALE). Walk-DMC has previously been *associated* with changes in gait measures after treatment, including overall gait deviation (GDI), consistent with the proposed causal diagram in this study^37^. The same study demonstrated that Walk-DMC was *associated* with changes in the sports and physical function sub-score of the pediatric outcomes data collection instrument (PODCI)^38^. While the PODCI sports and physical function score is not identical to the GMFM-66, it is reasonable to think that the two share a common underlying construct.

Static and dynamic motor control and are difficult to alter with treatments or therapies. Nevertheless, our results suggest that we should continue efforts towards finding control-enhancing therapies, as these hold the greatest prospect for impacting overall function. Spasticity is another common primary neurological impairment observed in children diagnosed with CP. Both the direct and total effects of spasticity were shown to be relatively small. Thus, intrathecal baclofen pump implantation, selective dorsal rhizotomy, or chemodenervation therapies aimed at spasticity reduction should not be expected to appreciably improve gross motor function. This prediction is consistent with what has been shown in outcome analyses of these treatments^10,39,40^.

Age did not have a significant effect. Thus, independent of neurological and orthopedic impairments, age alone is not expected to alter gross motor function substantially. In contrast, it has been previously shown that GMFM-66 improves from an early age and begins to plateau around age 8^41^. Approximately 85% of the individuals in this study were older than 7.5 (Table 1), and this may explain our finding that age was not an important cause of GMFM-66. In addition, the largest improvements due to ageing are expected in Gross Motor Function Classification (GMFCS)^42^ I individuals, who make up only 38% of this study population. Furthermore, we made the simplifying assumption of linearity, which cannot accurately model the growth and plateauing nature of previously observed GMFM-66 *vs*. age response. Lastly, the well-known GMFM-66 *vs*. age curves are stratified by GMFCS. While GMFCS *may* constitute a reasonable adjustment set, our causal model would suggest the nature of GMFM-66 changes are more complex. In fact, GMFCS level can be thought of as a causal consequence of many factors in our model. Put another way, children with poorer strength and motor control and worse spasticity are likely to be classified with a GMFCS level indicating greater severity of the disease. Thus, adjusting for GMFCS *alone* may not fully capture the effects that motor control, strength, orthopedic impairments, and gait impairments have on function. The lack of a direct age effect in our model suggests that changes in other factors with age (*e*.*g*. Ankle Dorsiflexion, Strength, Gait Pattern) may explain the apparent age effect observed in the GMFM-66.

The effect sizes of strength and gait quality (GDI) were both significant, suggesting that they are important mediators of function. Both variables can be improved by treatment (strengthening, physical therapy, orthopedic surgery). Our model suggests that improvement in these domains can raise gross motor function. However, the evidence for this is not clear. Outcome of these therapies have not consistently demonstrated improved function in patients^43^. One possible explanation could be the magnitude of strength or GDI improvement achieved with treatment. The standardized effect size for strength and GDI are 0.23 and 0.15, respectively. This means that for one standard deviation of strength improvement, we should expect 0.23 standard deviations of GMFM-66 improvement (2.8 points for the data in this study), and for one standard deviation of GDI improvement we should expect 0.15 standard deviations in GMFM-66 improvement (1.9 points). Typical GDI improvements following surgery are on the order of 7 points, which would correspond to *∼*1.3 point improvement on GMFM-66 scores, well below the reported minimally clinical important difference^44^.

The model results highlight the challenges faced for improving function in children with CP. The largest effect sizes are found in the static (SCALE) and dynamic (Walk-DMC) motor control measures; impairments for which there are currently no highly effective treatments. In contrast, spasticity, which we can treat via rhizotomy or baclofen pump, has a very small impact on gross motor function. Strength and gait quality both have significant, but modest (*∼*0.25), effect sizes. Additionally, improvements in these domains following treatment tend to be small (∼0.5 standard deviations or less), leading to minimal functional gains. If we hope to improve function through these domains, we need more effective treatments for strength and gait.

A causal diagram is a hypothesis. Since counterfactual observations (*e*.*g*. a version of a specific child with one clinical parameter altered) do not exist, causal diagrams can never be fully proven or falsified. They can, however, be shown to be reasonable from both a domain knowledge and statistical perspective. We showed that the proposed diagram is plausible and explained 75% of the variance in GMFM-66. Nevertheless, unobserved and unmeasured variables may play a significant role. Some potentially important factors were omitted, largely for practical purposes. Cognition may be one such omitted but important variable^45^. It is likely that cognition plays a meaningful role in function and including this variable could change the effect sizes of the factors we have examined. Socio-economic factors, access to resources, participation in the community, availability of recreational sports opportunities, emotional health, and myriad other complex factors could also be considered in future extensions of this model.

Similar to Kim and Park’s less comprehensive causal model of function^27^, we found that both spasticity and strength had significant effects on GMFM-66. However, the inclusion of additional causal factors significantly changed the absolute and relative magnitude of the effect sizes. Kim and Park estimated the effect sizes for spasticity and strength to be 0.34 and 0.45, respectively, while we found them to be 0.05 and 0.23. Differences in testing methods for these subjective measures may significantly impact absolute effect sizes, but should have less impact on relative effect sizes, which were also substantially different in our model compared to Kim and Park’s (4.82 and 1.32, respectively).

One limitation to this study was the exclusion of 14 individuals (4%) due to missing data. In some cases, data was not collected due to the patient’s inability to follow instructions. These missing data have potential to bias the results since some of the lowest functioning individuals may have been excluded. However, only six subjects (2%) were excluded for this reason. In addition, both sides of bilateral measures for unilateral individuals were included and averaged. While this may diminish the influence of the condition on the hemiparetic side, GMFM-66 is a score for an individual, allowing compensatory contributions of the non-hemiparetic side, and thus we reasoned that averaging sides was an appropriate approach.

This study should be viewed as a starting point, and not as a definitive model of causality and function in children with CP. We proposed a model, demonstrated its plausibility, and estimated the causal effect sizes of key clinical factors. We also showed important differences in interpretation that arise when causality, rather than simple association, is considered. The strength of our approach is the explicit proposal of a causal diagram, leading to *a priori* statistical models. This contrasts with much of the previous work in this area, which commonly examines *ad hoc* bivariate relationships in an unstructured methodology. In many cases, the bivariate approach leads to significant and strong correlations, but these findings may be misleading since they lack the proper causal context.

As noted earlier, lurking behind virtually every study of association is a hidden belief in a causal relationship. Except for purely predictive models, we suspect no researchers examine associations without some implied causality. While our proposed model is imperfect, it is transparent and explicit. Additional work is needed to discover more complete causal models and to devise experiments that confirm or refute the results of these models. The complexity of CP and the relationships among commonly measured variables demand that causality be considered when interpreting data.

## Methods

IRB approval was obtained for this retrospective analysis.

### Causal Model

A causal model^46^ was proposed to explain how a set of important, commonly measured, and potentially treatable neurological and orthopedic impairments affect function as measured by GMFM-66 (Table 3). The relationships in the model, along with linearity assumptions, lead to conditional independence tests that assess the plausibility of the model and covariate adjustment sets for statistical models used to estimate the relative causal contributions of various factors.

**Table 3.** Causal Model.

| Clinical Feature | Causes |
| --- | --- |
| Injury |  |
| Age |  |
| Motor Control | Injury |
| Static Motor Control | Motor Control, Age |
| Dynamic Motor Control | Motor Control, Age |
| Strength | Injury, Motor Control, Age* |
| Spasticity | Injury, Age |
| Ankle Dors. Contracture** | Strength, Spasticity, Dynamic & Static Motor Control, Age |
| Knee Ext. Contracture** | Strength, Spasticity, Dynamic & Static Motor Control, Age |
| Hamstrings Contracture** | Strength, Spasticity, Dynamic & Static Motor Control, Age |
| Hip Ext. Contracture** | Strength, Spasticity, Dynamic & Static Motor Control, Age |
| Tibial Torsion | Strength, Spasticity, Dynamic & Static Motor Control, Age |
| Femoral Anteversion | Strength, Spasticity, Dynamic & Static Motor Control, Age |
| Gait Pattern | Strength, Spasticity, Dynamic & Static Motor Control, Age,<br>Ankle ROM, Knee ROM, Popliteal ROM, Hip ROM,<br>Tibial Torsion, Femoral Anteversion |
| GMFM-66 | Strength, Spasticity, Dynamic & Static Motor Control, Age, Ankle ROM,<br>Knee ROM, Popliteal ROM, Hip ROM, Gait Pattern |

\* Because we measure strength in an age-adjusted manner, we do not include an explicit causal link in our graphical model used for testing plausibility and determining adjustment sets
\*\* ROM = Range of Motion. Ankle ROM, Knee ROM, Popliteal ROM, and Hip ROM also influence each other. This is modeled by all four ROM variables being caused by an unmeasured latent variable $U \rightarrow$ Ankle ROM, $U \rightarrow$ Knee ROM, $U \rightarrow$ Popliteal ROM, $U \rightarrow$ Hip ROM.

The causal model starts with the neurologic injury that has led to the diagnosis of CP (Table 3). The primary impairments resulting from this injury commonly manifest as spasticity, and reductions in static motor control, dynamic motor control, and strength. Each of these neurological factors is hypothesized to have a direct effect on gross motor function. These primary impairments affect gait through improper muscle activation patterns, which in turn affects GMFM-66. Static motor control also affects strength by impeding the ability to isolate and volitionally activate muscles, resulting in functional weakness. Strength in turn affects both gait and gross motor function. The effect of age on gross motor function is accounted for both indirectly, through neuromaturation and growth, and directly, which reflects motor learning associated with practice. Orthopedic deformities, in the form of contractures (Ankle, Knee, Hamstrings, and Hip) are partly caused by the improper muscle forces brought about by spasticity, poor strength, and poor motor control. These improper forces also influence the remodeling of bony torsions (Femoral Anteversion and Tibial Torsion). The contractures and long-bone torsions both affect gait, and the contractures impede gross motor function. Note that we assume no measurable effect of torsions on gross motor function as measured by the GMFM-66. There are undoubtedly other impairments associated with the injury that are not represented. For the present model these other factors are ignored. This meaningful limitation and its impact on our results will be discussed later.

### Study Data

We evaluated the model using 300 out of 314 consecutive unique individuals with a diagnosis of CP who were seen at a single center for a quantitative 3D gait assessment including electromyography (EMG) of the lower extremity. A query of the laboratory database created a dataset for these individuals which included CP subtype classification (bilateral or unilateral), gross motor function (GMFM-66), age at assessment (Age), walking dynamic motor control (Walk-DMC^28^), static selective motor control total limb scores (SCALE^29^), gait impairments (gait deviation index or GDI^31^), and muscle strength (manual muscle testing grade and hand-held dynamometry^30,47^). Physical exam measures of limb rotational deformities (femoral anteversion, tibial torsion) and sagittal plane joint muscle contractures (popliteal angle, maximum hip extension, knee extension, and ankle dorsiflexion) were also included and expressed as differences from levels of typically developing individuals. Only subjects with complete datasets were included. For all bilateral measures, averages between sides were used, including for unilaterally involved individuals.

Dynamic motor control was expressed by the Walk-DMC measure, calculated from electromyography collected during walking at self-selected speed. Walk-DMC is a validated measure of motor control that is scaled so that 100 (10) is the mean (standard deviation) of typically developing peers. Walk-DMC values were computed based on the methods described by Steele *et al*.^28^, using an aggregate analysis of 5-7 self-selected speed trials obtained as part of the clinical gait analysis protocol.

Static selective motor control was measured using the SCALE as evaluated by an experienced physical therapist. Total limb SCALE values (0-10 scale) were used as an overall representation of static motor control^29^.

Spasticity was evaluated on a 0-5 scale derived from multiple measures collected during physical examination by an experienced physical therapist^34^. Briefly, the spasticity scale included modified Ashworth scales^48^ recorded for between 3 to 5 muscles, Duncan-Ely tests of rectus femoris spasticity, beats of clonus, and ankle plantarflexor differences between initial catch of quick stretch and maximal values.

Strength was measured by both subjective evaluation of muscle grades and handheld dynamometry. At our center, muscle strength grades are routinely recorded for abdominals, hip flexors, extensors, abductors and adductors, knee flexors and extensors, and ankle flexors and extensors^30^. Hand-held dynamometry is routinely performed for knee and ankle flexors and extensors on those individuals with adequate strength and ability to follow instructions^47^. After adjusting for age, gender, and weight these values are expressed as Z-scores relative to typically developing controls collected^49^. When dynamometry is collected, subjective muscle strength grades are not recorded for these muscles. In order to compile the most comprehensive and consistent data set, and to ensure that weaker subjects were not systematically excluded, dynamometry values were converted into a muscle grade using Z-scores (Z>-1 = grade 5, -2<Z<- 1 = grade 4+, -3<Z<-2 = grade 4, -4<Z<-3 = grade 4-, Z<-4 = grade 3+). All subjective muscle grades were assigned numerical values with +/-ratings decremented in thirds (*e*.*g*. grade 4- = 3.67, grade 4 = 4.0, grade 4+ = 4.33). Grades were then averaged on a 0-5 scale across the 8 bilateral muscle groups plus abdominals.

Femoral anteversion was estimated from the difference between maximum inward and outward rotation of the hip with the subject prone and knee flexed to 90^35^. Tibial torsion was estimated using bimalleolar and epicondyle landmarks with the subject supine and the knee extended^32,33^. Ranges of motion were measured with handheld goniometry. Hip extension contracture was measured with the opposite hip flexed, knee extension contracture in a supine position, hamstrings contracture from the popliteal angle taken with the opposite leg extended, and ankle dorsiflexion contracture in a prone position with the knee at maximal extension^32,33^. Gait quality was measured by the gait deviation index (GDI), computed using published techniques^31^. The GDI is a validated measure of overall gait deviations, scaled so that 100 (10) is the mean (sd) for typically developing controls.

In all models, variables were transformed as follows. First, each variable was standardized using the sample mean and standard deviation. Next, the sign of the variable was adjusted so that larger values indicate less impairment and smaller values indicate more impairment. This was done so that positive effect sizes imply a higher score (better function) on the GMFM-66 scale. For age, larger values retain the conventional meaning of older.

## Analysis

The conditional independencies are a list of partial correlations that must be near zero or insignificant for the observed data to be consistent with the structure of the hypothesized causal model^50^. Implied conditional independencies were identified to test the plausibility of the proposed model^46^. Meeting these conditions is necessary, but not sufficient, for a causal model to be correct, since there can be multiple models that meet the same implied independencies.

We chose to use linear models to estimate the magnitude and sign of total and direct causal effects for each factor of interest. A linear model was built for each factor whose effect size was being investigated using the adjustment set identified from the causal model. Adjustment sets are lists of covariates that, when included in the linear model, allow the causal effect of a particular factor to be estimated by controlling appropriately for other factors. Only main effects were allowed in the linear models (no interactions). This decision was driven by both causal and practical considerations. From a causal perspective, we did not see clear rationale for interaction effects, and, if they did exist, we assumed they would likely be significantly smaller than the main effects. From a practical perspective, we know that spurious significant interactions are likely, given the number of factors under consideration, and that approximately 16x more samples are needed to detect significant interactions compared to main effects^51^.

Thus, in light of our relatively coarse measurements and modest sample size, we settled on a main-effects-only model. In addition to deriving causal effect sizes for each *individual* factor, we also built a predictive model to examine the total causal effect of the factors taken together. The predictive model allows us to estimate GMFM-66 as a function of the fundamental clinical measures of neurological and orthopedic impairment.

We also derived bivariate linear models to assess the apparent, non-causal, effect sizes for each of the factors considered in the causal model. For example, we regressed GMFM-66 against Strength as a single predictor, then against Spasticity as a single predictor, and so on. The effect sizes from the bivariate models typify the naïve estimates that result from the non-causal association analyses that pervade the literature. Thus, comparisons between the two approaches illustrate the interpretive differences that arise as a result of the causal approach.

All modeling and statistics were performed with R (version 4.02, R Core Team, 2020). Causal model building and testing was done using dagitty^46^.

## Data Availability

Our institution does not allow us to share data due to privacy concerns.

